# The utility of MRI radiological biomarkers in determining intracranial pressure

**DOI:** 10.1101/2023.04.26.23289164

**Authors:** Anand S. Pandit, Musa China, Raunak Jain, Arif H.B. Jalal, Shivani.B. Joshi, Crystallynn Skye, Zakee Abdi, Yousif Aldabbagh, Mohammad Alradhawi, Ptolemy D. W. Banks, Martyna K. Stasiak, Emily B.C. Tan, Fleur C. Yildirim, James K. Ruffle, Linda D’Antona, Hasan Asif, Lewis Thorne, Laurence D. Watkins, Parashkev Nachev, Ahmed K Toma

**Affiliations:** High-Dimensional Neurology Group, UCL Queen Square Institute of Neurology, University College London, London, UK; Victor Horsley Department of Neurosurgery, National Hospital for Neurology and Neurosurgery, London, UK; University College London, UCL, Division of Medicine, London, United Kingdom; Department of Psychology and Language Sciences, University College London, London, United Kingdom; School of Medicine, Barts and The London School of Medicine and Dentistry, Queen Mary University of London, London, UK

**Keywords:** adult hydrocephalus, Bayesian modelling, radiological biomarkers

## Abstract

**Background:** Intracranial pressure (ICP) is a physiological parameter that conventionally requires invasive monitoring for accurate measurement. Utilising multivariate predictive models, we sought to evaluate the utility of non-invasive, accessible MRI biomarkers in predicting ICP and their reversibility following cerebrospinal fluid (CSF) diversion.

**Methods:** The retrospective study included 325 adult patients with suspected CSF dynamic disorders who underwent brain MRI scans within three months of elective 24-hour ICP monitoring. Five MRI biomarkers were assessed: Yuh sella grade, optic nerve vertical tortuosity (VT), optic nerve sheath distension (ONSD), posterior globe flattening (PGF) and optic disc protrusion (ODP). The association between MRI biomarkers and 24-hour ICP was examined and reversibility of each following CSF diversion was assessed using uni- and multivariate techniques.

**Results:** All five biomarkers were significantly associated with median 24-hour ICP (p<0.0001). Using a pair-wise approach, the presence of each abnormal biomarker was significantly associated with higher median 24- hour ICP (p<0.0001). On multivariate analysis, ICP was significantly and positively associated with Yuh grade (p<0.0001), VT (p<0.0001) and ODP (p=0.003), after accounting for age and suspected diagnosis. Bayesian multiple linear regression predicted 24-hour median ICP with a mean absolute error of 2.71 mmHg. Following CSF diversion, we found Yuh grade to show significant pairwise reversibility (p<0.001).

**Conclusions:** ICP was predicted with clinically useful precision utilising a compact Bayesian model, offering an easily interpretable tool that utilised non-invasive imaging data. MRI biomarkers are anticipated to play a more significant role in the screening, triaging, and referral of patients with suspected CSF dynamic disorders.

**Key messages:**

- Brain MRI biomarkers have been found to be correlated with cerebrospinal fluid (CSF) pressures. However, previous studies have not examined these imaging features with continuous intracranial pressure (ICP) measurements, or in patient cohorts with sizable numbers or different CSF dynamic disorders than idiopathic intracranial hypertension.
- In this retrospective cohort study, patients with abnormal neuroradiological markers (optic nerve sheath diameter, pituitary: sella grade, optic nerve vertical tortuosity, posterior globe flattening or optic disc protrusion) had significantly higher median 24-hour ICP readings. After adjusting for age and diagnosis, Yuh sella grade, vertical tortuosity and optic disc protrusion were significantly associated with ICP. Our multiple linear regression model was able to predict 24-hour median ICP using routine MR-imaging in those with chronic CSF disorders. Pituitary deformation resolved following CSF diversion, suggesting reversibility of certain radiological biomarkers.
- Brain MRIs are widely accessible and non-invasive, and are commonly used in elective patients with suspected raised ICP. Our study provides a tool incorporating simple clinico-radiological parameters for the screening, triaging, and referral of patients with suspected abnormal ICP, and our results have important implications for the diagnostic routine of patients with suspected intracranial hypertension.

## Introduction

Abnormal intracranial pressure (ICP) represents an important pathophysiological parameter that portends neurological harm. Raised ICP can manifest as an acute phenomenon in patients following trauma and in those with intracranial space-occupying lesions. It is also apparent in patients whose disease process is more insidious and without an underlying structural cause as in the case of idiopathic intracranial hypertension (IIH) [1,2].

Invasive ICP measurement via intraparenchymal monitoring or external ventricular drain remains the gold standard method of continuous measurement of ICP [2,3]. Whilst direct monitoring tools provide accurate, continuous real-time measurements of ICP, their invasive nature carries surgical risks and requirements for inpatient hospitalisation: limiting their use to tertiary-level centres with a dedicated neurosurgical service. On the other hand, lumbar puncture and the use of manometry provide a ‘snapshot’ estimate of ICP that is susceptible to inaccuracy, dependent on patient positioning, natural physiological and diurnal variations [4,5], and is not entirely free from procedural risk.

Much desired are assessments of ICP that are non-invasive and can guide the management of patients with suspected abnormalities related to cerebro-spinal fluid (CSF) dynamics. Ophthalmological markers such as changes to the optic disc, lack of spontaneous venous pulsation and ultrasound markers of an enlarged optic nerve sheath show promise in predicting raised ICP [6,7]. However, there are practical considerations regarding availability and training, and it remains unclear the degree to which these markers are sensitive [8].

Brain MRIs are widely accessible in non-emergency clinical settings and are commonly performed in elective patients with suspected raised ICP. In addition to pituitary gland shape, MRI biomarkers such as distension of the optic nerve sheath, flattening of the posterior globe, tortuosity and protrusion of the optic nerve have been posited as proxy indicators of raised ICP in patients with IIH [9–15]. However, few studies correlate these imaging features with ICP measurements, nor have been examined in patient cohorts with sizable numbers or with cerebrospinal fluid abnormalities other than IIH. Others have utilised these biomarkers to predict ICP, dividing patient cohorts in a dichotomous manner as per arbitrary thresholds for raised ICP, which are likely to be contentious and to vary across different patient cohorts [15].

To that end, we retrospectively evaluate the utility of common MR-imaging biomarkers in prediction of intracranial pressure in the largest cohort of neurological and neurosurgical patients admitted for 24-hour intracranial pressure recording available. Each biomarker is assessed individually and in the context of both classical and Bayesian multivariate models in order to provide estimates of certainty on predictions. As a secondary aim, in patients who have had CSF diversion and attempted normalisation of ICP, we determine whether these biomarkers are reversible.

## Methods

### Ethics and guidelines

This use of data from this cohort was approved by the North East-Newcastle & North Tyneside 2 Research Ethics Committee and the Health Research Authority (20/NE/0127). Written consent was waived due to the retrospective nature of the study. All patients consented to ICP monitoring. The study follows the TRIPOD checklist for multivariate predictive models where relevant.

### Patients

A retrospective review of our institutional database was conducted to identify a consecutive series of all adult patients investigated with elective 24-hour intraparenchymal ICP monitoring between January 2006 and December 2021 at a large tertiary neurosciences centre. Patients meeting the following eligibility criteria were selected: (1) they had undergone elective 24-hour ICP monitoring for a suspected CSF dynamic disorder and (2) had a brain MRI performed within three months of ICP monitoring, following a protocol described in [15]. Patients who had CSF diversion prior to the monitoring were also included. Unless otherwise specified, all patient diagnoses were made by a consultant neurologist or neurosurgeon following evaluation of the combined clinical, imaging and ophthalmological information.

### ICP Monitoring

Within our institution, patients with suspected CSF dynamics abnormalities undergo elective ICP monitoring often after discussion in a multidisciplinary team meeting consisting of consultant neurosurgeons and neurologists with a specialist interest in hydrocephalus and CSF disorders. A detailed description of the standardised protocol for the elective 24-hour ICP monitoring procedure at our institution has previously been published [16]. Our standard operating procedure involves insertion of a right frontal Raumedic™ intraparenchymal ICP probe under local anaesthesia with or without sedation in the operating theatre. For a continuous period of 24 hours following insertion, high-frequency ICP data (100 Hz) is collected, and patients are encouraged to mobilise to simulate their typical level of activity. Monitoring ceases after the 24-hour period or is extended until the attending neurosurgeon (with a specialist interest in adult CSF disorders) deems the ICP monitoring data to be of sufficient quality. The raw data is downloaded and then analysed using the ICM+ software (University of Cambridge, UK), into minute-by-minute ICP values and amplitude. These are further processed yielding a number of ICP parameters over the entire 24 hours of which median ICP, the most frequently used parameter was chosen as the outcome variable.

### Neuroradiology

Diagnostic brain MRI scans performed within three months of the elective 24-hour ICP monitoring were selected. Although the majority were performed internally at our centre, external transferred scans were used if these were not available.

T1-weighted sagittal brain MRI sequences were used to grade pituitary height and optic nerve vertical tortuosity. Maximum pituitary gland height was measured on the mid-sagittal T1w scans and graded from 1 to 5 according to the Yuh et al. 2000 classification [12]. A pituitary gland height of less than one-third of the height of the sella turcica is considered abnormal; this corresponds to grades 3-5 of the Yuh classification system (Figure 1A). Optic nerve vertical tortuosity (VT) was confirmed on the appearance of an “s-shaped’’ or “kinked” appearance of the optic nerve on T1w sagittal sequence MRI (Figure 2A) [9]. T2-weighted axial brain MRI sequences (non-fat-suppressed) were used to grade optic nerve sheath distension (ONSD), optic disc protrusion (ODP) and posterior globe flattening (PGF). ONSD was measured in an axis perpendicular to the optic nerve, measuring from the outer margin of the optic nerve to the outer margin of the optic nerve sheath. ONSD was confirmed when the measurement of the hyper-intense CSF-containing sheath surrounding the optic nerve was more than 2 mm in width at any point up to 10 mm behind the globe (Figure 2C) [9]. Optic disc protrusion was confirmed, where at the point of attachment of the optic nerve, there was intraocular protrusion of the optic disc and associated concavity of the sclera (Figure 2G) [9,14]. VT, ONSD, ODP and PGF were graded between 0 to 2 depending on whether they had demonstrated this sign in none, one or both eyes. An abnormal grade was considered if at least one optic nerve was affected.

**Figure 1.**
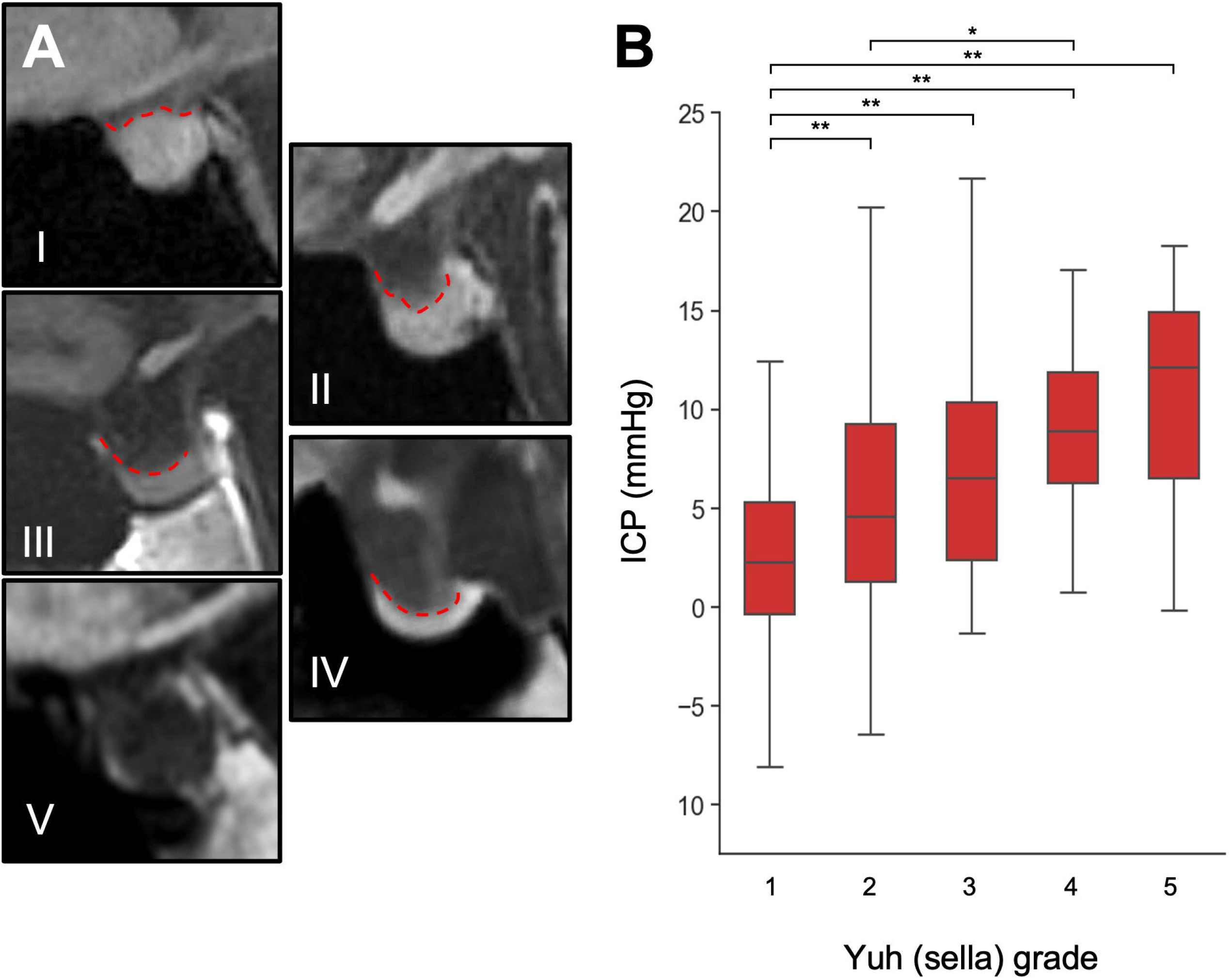
Yuh sella grading in selected patient examples using T1-weighted sagittal images (A), and association with 24-hour median ICP (B). (* p <0.05; ** p <0.01; *** p <0.001)

**Figure 2.**
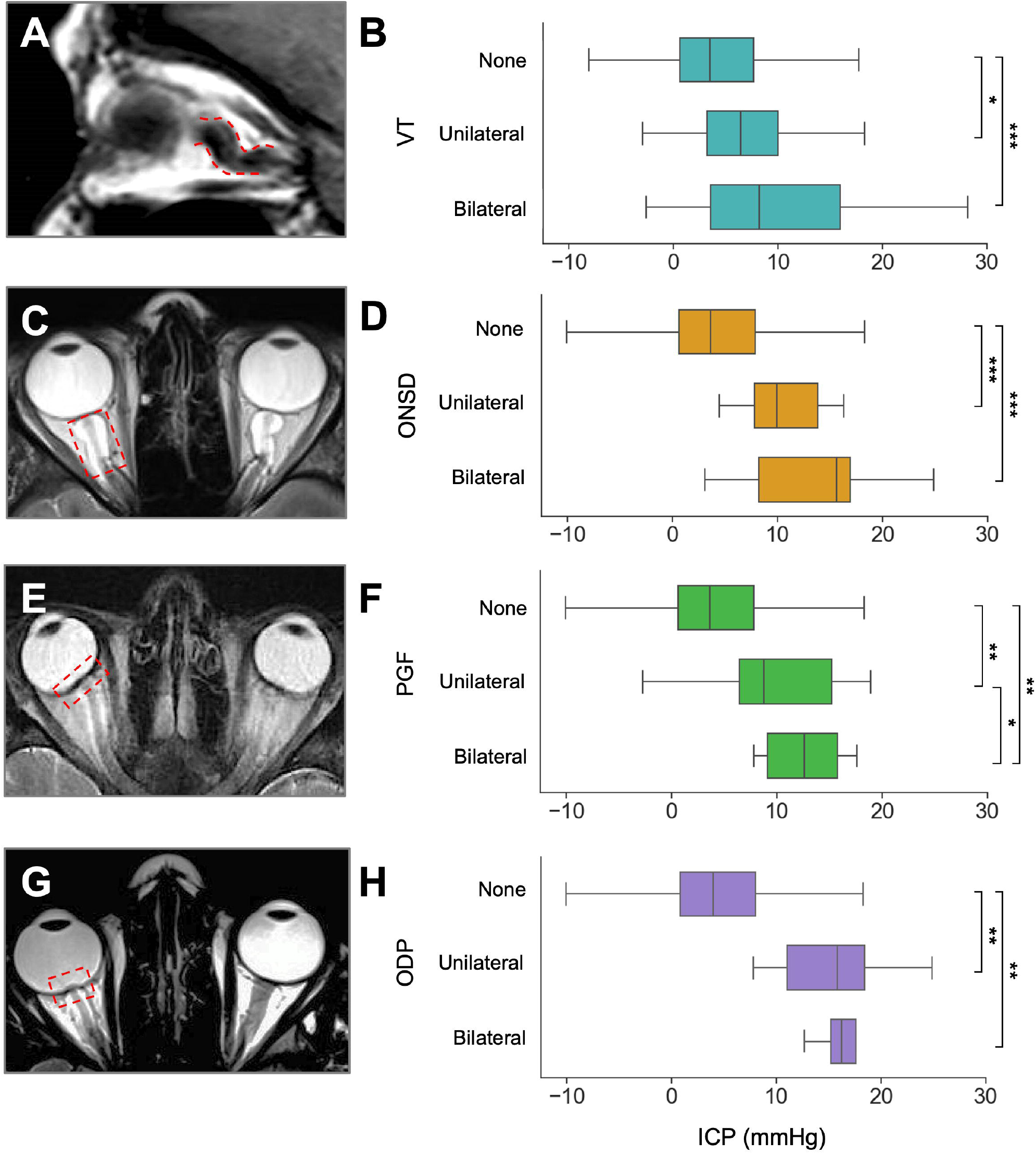
Non-pituitary MRI markers in selected patient examples using axial T2-weighted imaging and associated 24-hour median ICP measurements for optic nerve vertical tortuosity (A, B); optic nerve sheath distension (C, D); posterior globe flattening (E, F); optic disc protrusion (G, H)

Biomarker assessment was performed in several stages to assure inter-rater reliability and were independently graded by a board-certified neuroradiologist blinded to clinical and ICP information with moderate to high inter-rater reliability (see Supplementary Methods).

### Data analysis

An extensive exploratory data analysis was first performed, investigating the distribution of both explanatory and target variables. All statistical analyses were performed in Python (v=3.8.2).

Univariate testing examined the association between individual biomarkers and median 24-hour ICP. Group averages were compared using either a two-sided independent samples t-test or Mann-Whitney U (MWU) and Kruskal-Wallis (KW) test depending on whether the data was parametric. Normality of data was assessed by histogram visualisation and using the Kolmogorov-Smirnov test. Both raw and binarised (normal/abnormal) grading were evaluated. Significant results were reported if p was <0.05 following multiple comparison correction using the Holm-Bonferroni method. Reversibility testing of each radiological biomarker was assessed using either a paired t-test or Wilcoxon-Sign-Rank test in patients who were surgically-naive and then underwent CSF diversion.

Multivariate testing was first performed using an ordinary least squares (OLS) regression examining the association between radiological biomarkers and ICP, with and without adjustment for clinical and demographic variables. Here, model assumptions were verified including assessment of homoscedasticity and normality among the residuals. Next, a Bayesian multiple linear regression model was fitted [17]. Bayesian methods offer several benefits relative to classical methods including: (i) flexibility when fitting complex and realistic models, (ii) incorporation of prior information about plausible values for the model parameters, and (iii) the outputs, which as expressed in terms of probabilities are generally easier to interpret by non-expert users. Overall model performance was evaluated using standardised scores, namely adjusted R^2^ and mean absolute error based on leave-one-out cross validation [18] (see Supplementary Methods for details regarding model implementation).

## Results

### Patient characteristics

325 patients (229 female) met our participant criteria with a mean age of 40 years (SD = 15.1) [Table 1]. The most frequent diagnosis or suspected diagnosis prior to ICP monitoring was IIH (30%), followed by patients with syringomyelia or a Chiari malformation (19%). 65 patients (20%) had no formal diagnosis or underlying structural abnormality and were being investigated for persistent, non-specific high or low intracranial pressure symptoms.

**Table 1.** Summary of patient demographics, underlying conditions and interventions. CFO = cyst fenestration with Ommaya reservoir insertion; CPS = cysto-peritoneal shunt; ETV = endoscopic third ventriculostomy; EVD = external ventricular drain; FMD = foramen magnum decompression; ICPM = intracranial pressure monitoring; IIH = idiopathic intracranial hypertension; IQR = interquartile range; LD = lumbar drain; LOVA = long standing overt ventriculomegaly in adults; LPS = lumboperitoneal shunt; NPH = normal pressure hydrocephalus; SOL = space-occupying lesion; SPS = syringo-pleural shunt; VAS = ventriculo-atrial shunt; VPS = ventriculoperitoneal shunt; VSS = venous sinus stenting.

|  |  |
| --- | --- |
| **Patients** | 325 |
| **Mean age (years)** | 40.1 +/- 15.1 |
| **Gender (F/M [%])** | 229/96 [70.5/29.5] |
| **Primary pathology (%)** | IIH = 97 (29.8)<br>Chiari-Syringomyelia = 61 (18.8)<br>Tumour / SOL = 32 (9.8)<br>Congenital = 18 (5.5)<br>NPH = 15 (4.6)<br>Aqueductal Stenosis = 13 (4.0)<br>LOVA = 10 (3.1)<br>CSF leak = 8 (2.5)<br>Neurovascular = 3 (0.9)<br>Traumatic brain injury = 3 (0.9)<br>Non-specific = 65 (20.0) |
| **Median days between ICPM and MRI (IQR)** | 31 (7-60) |
| **Prior intervention (%)** | None = 181 (55.7)<br>VPS = 88 (27.1)<br>LPS = 17 (5.2)<br>FMD = 6 (1.8)<br>VSS = 5 (1.5)<br>ETV = 3 (0.9)<br>CFO = 3 (0.9)<br>VAS = 2 (0.6)<br>CPS = 2 (0.6)<br>EVD = 2 (0.6)<br>Multiple = 16 (4.9) |
| **Surgical intervention following ICP / MRI (%)** | No further surgical intervention = 239 (73.5)<br>VPS = 59 (18.1) |
|  | LPS = 6 (1.8)<br>LD = 5 (1.5)<br>FMD = 5 (1.5)<br>CFO = 3 (0.9)<br>VSS = 2 (0.6)<br>EVD = 1 (0.3)<br>VAS = 1 (0.3)<br>SPS = 1 (0.3)<br>Multiple = 3 (0.9) |

The median time between ICP monitoring and MRI scan was 31 days. No correlation was found between median ICP and the MRI time interval. 144 patients had a prior CSF-diverting intervention, of which the majority had insertion of a ventriculo-peritoneal shunt. No significant difference was found in intracranial pressure between patients with and without prior CSF diversion.

### The association of individual radiological biomarkers with intracranial pressure

#### Sella grade

Higher Yuh sella grade (more empty sella) was associated with a higher intracranial pressure (Kruskal-Wallis statistic = 57.2, p < 0.0001), with a stepwise increase in ICP between each grade (Figure 1B). Across these increases, significant differences were found between sella grade I and all other grades and between sella grade II and IV. Using a binarised approach, abnormal sella morphology (Yuh grade 3-5) was linked with higher ICP (9.5 mmHg) as compared to those with ‘normal’ sella shape (Yuh grades 1-2, 3.5 mmHg) [Mann-Whitney U = 8596, p <0.0001].

#### Vertical tortuosity

Patients with tortuous optic nerves had a higher intracranial pressure (KW = 17.2, p < 0.001), with a significant pairwise increase between none and unilateral and none and bilateral signs (Figure 2B). Using a binarised approach, patients who had either one or two tortuous optic nerves had a significantly higher median ICP (6.7 mmHg) as compared to those without signs of tortuosity (3.5 mmHg, MWU = 12358, p < 0.0001).

#### Optic nerve sheath distension

Patients with a distended optic nerve sheath had a higher intracranial pressure (KW = 29.4, p < 0.0001), with a significant stepwise increase between those with no ONSD and unilateral ONSD or bilateral ONSD (Figure 3B). Using a binarised approach, patients who had either one or two optic nerves with nerve sheath distension had a significantly higher median ICP (10.1 mmHg) as compared to those without signs (3.7 mmHg, MWU = 5605.5, p < 0.0001).

**Figure 3.**
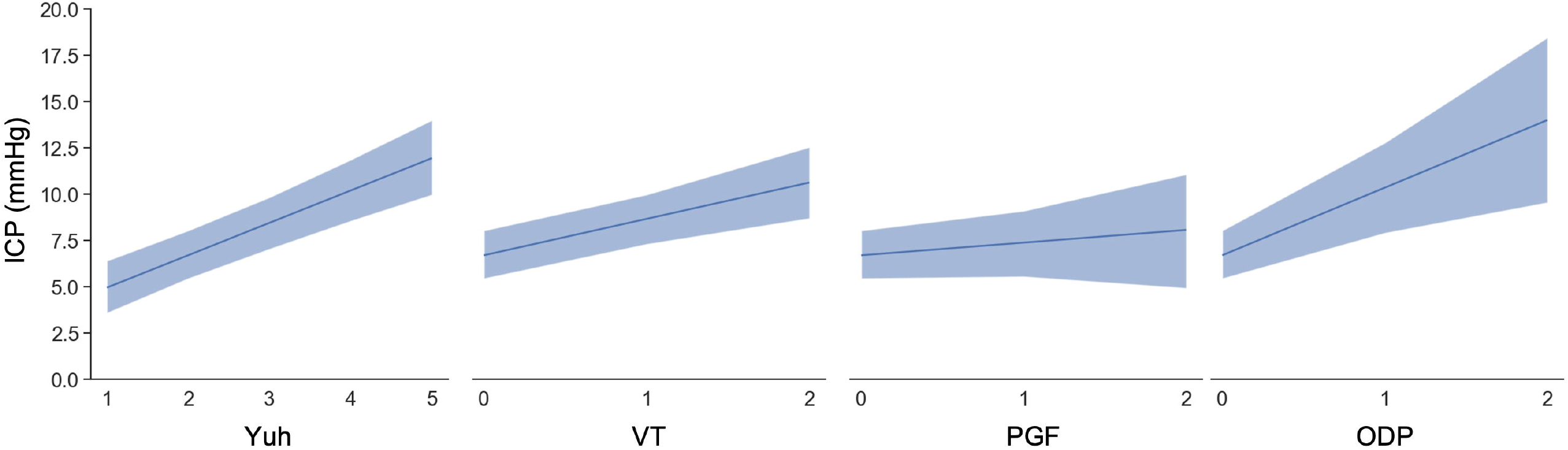
Conditional adjusted predictions for individual radiological biomarkers. All other covariates, including other radiological markers, are constant at their sample mean. Dark blue line represents mean with 94% credible intervals shown in the pale blue bars. ODP = optic disc protrusion, PGF = posterior globe flattening, VT = vertical tortuosity, Yuh = pituitary sella grade.

#### Globe flattening

Patients with optic globe flattening had a higher intracranial pressure (KW= 30.2, p < 0.0001), with a significant pairwise increase between none, unilateral and bilateral signs (Figure 4B). Using a binarised approach, patients who had either one or two optic globes showing flattened appearances had significantly raised ICP (10.1 mmHg) as compared to those without signs (3.7 mmHg, MWU = 6839.5, p < 0.0001).

#### Optic disc protrusion

Patients with protruded optic discs had a higher intracranial pressure (KW = 21.1, p < 0.0001), with a significant pairwise increase between none and unilateral and none and bilateral signs (Figure 5B). Using a binarised approach, patients who had evidence of either uni-or bilateral optic disc protrusion had a significantly raised median ICP (16.2 mmHg) as compared to those without signs (4.0 mmHg, Mann-Whitney U = 3278, p < 0.0001).

#### Reversibility

36 patients (28 female) met our criteria for subgroup reversibility analysis with a mean age of 40 years (SD = 18.0). The most frequent diagnosis or suspected diagnosis prior to ICP monitoring and intervention was Chiari malformation (17%) followed by IIH (14%). Four patients (11%) had no formal diagnosis or underlying structural abnormality and were being investigated for persistent, non-specific high or low intracranial pressure symptoms. No significant differences were found in this smaller data-set as compared to the remainder of the cohort with respect to age, sex or diagnosis. In this group, the median interval between intervention and post-intervention MRI scan was 235 days (IQR: 74 -529).

Among patients who were surgically naive and then who had CSF diverting treatment following ICP monitoring, only the sella grade was found to be reversible (pre-intervention Yuh median = 2 (IQR: 1-3), post-intervention Yuh median = 1 (IQR: 1-2), Wilcoxon Sign-Rank, p < 0.001) [Supplementary Table 1]. Among the remaining markers, ONSD demonstrated a trend toward reversibility (p < 0.1), although this value was uncorrected.

#### Multivariate prediction of ICP using radiological markers

Three multivariate models were fitted to best characterise the association between radiological markers and intracranial pressure.

In the first, a sparse ordinary least squares (OLS) model was fitted where radiological markers were used in the absence of any other information, to replicate the scenario if a clinician was reviewing a patient’s MRI scan blind to other clinical details as may be realistic. Given the high anticipated degree of collinearity between radiological markers, pairwise Cramér’s V associations were calculated and the range of ICPs for patients with abnormal markers were delineated (Supplementary Figure 1). As there was correlation and overlap between optic nerve sheath distension and globe flattening, the former was removed from the model, resulting in no change in adjusted R^2^. This model explained approximately a quarter of the variance (adjusted R^2^ = 0.26), and all four remaining radiological markers were significantly, positively, and independently associated with ICP (Table 3). This model outperformed univariate OLS regression for each radiological marker.

**Table 3.** Comparison of multivariate models used to predict 24-hour median intracranial pressure. CI = confidence interval; CrI = credible interval; LOO-CV = leave-one-out cross validation; ns = non-significant; ODP = optic disc protrusion, OLS = ordinary least squares; ONSD = optic nerve sheath diameter; PGF = posterior globe flattening; VT = vertical tortuosity; Yuh = pituitary sella grade. (* see Supplementary Table 2; ** see Supplementary Table 3)

| Model |  | Radiological (univariate) |  | Radiological (multivariate) |  | Clinico-radiological |  | Clinico-radiological |
| --- | --- | --- | --- | --- | --- | --- | --- | --- |
| Method |  | OLS |  | OLS |  | OLS |  | Bayesian linear |
| Adjusted R<sup>2</sup> |  | 0.03 - 0.10 |  | 0.26 |  | 0.33 |  | 0.37 |
| LOO-CV<br>Mean Absolute Error (mmHg) |  | 3.83 - 4.00 |  | 3.20 |  | 2.83 |  | 2.71 |
| Variable |  | Coefficient (CI) | p | Coefficient (CI) | p | Coefficient (CI) | p | Mean coefficient (94% CrI) |
| Clinical | Age |  |  |  |  | - 0.07 (-0.12 – -0.03) | 0.003 | -0.07 (-0.11 – -0.03) |
|  | Diagnosis |  |  |  |  | *variable | - | **variable |
| Radiological | Yuh | 2.25 | <0.0001 | 1.77 (1.19 - 2.36) | <0.0001 | 1.76 (1.19 - 2.32) | <0.0001 | 1.76 (1.19 - 2.28) |
|  | VT | 2.74 | <0.0001 | 2.03 (1.04 - 3.03) | <0.0001 | 1.96 (0.99 - 2.92) | <0.0001 | 1.95 (1.05 - 2.88) |
|  | PGF | 4.27 | <0.0001 | 1.74 (0.05 - 3.42) | 0.04 | 0.69 (-0.95 - 2.33) | ns | 0.70 (-0.83 - 2.32) |
|  | ODP | 7.28 | <0.0001 | 4.57 (2.09 - 7.05) | <0.0001 | 3.62 (1.22 - 6.03) | 0.003 | 3.61 (1.33 - 5.89) |
|  | ONSD | 4.69 | <0.0001 |  |  |  |  |  |
| Intercept |  | - | - | 0.68 (-0.64 - 2.00) | ns | 3.51 (-0.11 - 7.12) | 0.06 | 3.49 (-0.03 - 6.94) |

In the second, a more complete OLS model was fitted that included salient demographic and clinical variables including patient age, sex, suspected diagnosis and whether they had a previous surgical intervention for CSF diversion. Variables were permuted and the model which minimised the Bayesian Information Criterion the most was selected (i.e., the most parsimonious model). In this way, patient age and diagnosis were retained. This model explained around a third of the variance (adjusted R^2^ = 0.33). Here, sella grade, VT and ODP remained significant after clinical confounder adjustment (Table 3). Individual diagnoses were not significantly associated with ICP after multivariate adjustment; however age did remain significant.

In the third, a Bayesian linear model (see Supplementary Material) was fitted using the previous clinical and radiological variables, however here no specification was made on the relationship between independent variables and median ICP. This had a higher adjusted R^2^ (0.37) as compared to the previous model, and one could assess the degree of credibility in the coefficients of each imaging variable when other variables were held at the sample mean (Figure 3). In all three models the use of multiplicative interaction terms showed no improvement in model scoring and are reported without interaction.

## Discussion

### Summary of results

This observational study investigates the association between five conventionally acquired MRI biomarkers and 24-hour median intracranial pressure in a cohort of 325 neurological and neurosurgical patients admitted for intracranial pressure monitoring. We found that patients exhibiting abnormalities in each radiological biomarker had significantly higher median 24-hour median ICP readings when assessed using univariate tests. After accounting for patient age and diagnosis in multivariate analyses, both frequentist and Bayesian approaches demonstrate significant, independent and positive associations between Yuh sella grading, vertical tortuosity and optic disc protrusion and ICP. Further, the Bayesian multiple linear regression model demonstrated the best performance, with the ability to robustly predict 24-hour median ICP with a cross-validated mean absolute error of only 2.71 mmHg: representing a clinically acceptable value for practical application. To that end, we derive a compact model for clinical use that can estimate ICP using routine MR imaging in those with chronic CSF disorders (Table 3, Supplementary Table 3). Finally we note that following CSF diversion in surgically-naive patients, there appears to be resolution of pituitary deformation as measured by improvements in Yuh sella grading, recapitulating posited reversibility of certain imaging biomarkers.

### Interpretation of results and context

Accurate, non-invasive prediction of ICP remains a much-desired tool for selected cohorts of patients suspected of having an underlying CSF-disorder. This faculty, if available, would help screen, triage and refer neurological and neurosurgical patients with abnormal ICP with greater precision. While clinical signs associated with high or low ICP have historically been well described, these may be ambiguous and non-specific [19]. On the other hand, ophthalmological and ultrasound-based markers have gained recent recognition but may not be widely available outside of specialised centres. In contrast, conventional MRI sequences are routinely obtained and offer a means to assess intracranial pressure by virtue of its morphological effects on the optic nerve, globe and sella. Previous studies utilising MR imaging biomarkers have been limited to small cohorts of patients, typically with IIH and may be correlated with non-standardised measurements of ICP [9,12,20–22]. Others divide patient cohorts in a dichotomous manner using arbitrary thresholds for raised ICP, which are likely contentious and variable across different patient cohorts [15,23]. Yet in spite of these differences, our univariate results remain broadly in tandem with the existing literature.

In a cohort of IIH patients, Rohr et al found that the number of radiographic MR imaging signs were positively associated with lumbar puncture (LP) opening pressure, whereas Tuncel et al found no such correlation [21,22]. These differences may be, at least in part, related to how each imaging marker was weighted and that only a single time point LP measurement was used to assess ICP [24]. Using 24-hour intraparenchymal ICP readings, D’Antona et al. investigated the association between pituitary gland shape, ONSD, VT, and ODP and median ICP within a cohort of 45 patients with a wide range of CSF dynamic disorders [15]. They found all four biomarkers, when abnormal, were significantly associated with raised 24-hour ICP readings and in 94% of patients in whom all four radiological signs were absent exhibited ‘normal’ median ICP readings (defined as <5.96 mmHg). Similarly with the addition of PGF, we found that when abnormal these radiographic markers were significantly associated with increased 24-hour median ICP on univariate analyses, and were associated with an even higher ICP if the marker was present bilaterally. Like D’Antona et al, we note that the ICP range associated with each abnormal radiographic marker is somewhat unique (Supplementary Figure 1), possibly reflecting the differential spatial effects of intracranial pressure as it rises beyond normal limits. Caution is warranted, however, on placing sole emphasis on any single MRI imaging sign to inform clinical decisions. Chen et al. suggested that certain abnormal MRI signs are also prevalent in patients investigated for non-CSF dynamic disorders [25]. Approximately a third of patients who had mixed indications for scanning, in their study had an ‘empty sella’ sign and up to 11% had increased perioptic CSF suggestive of ONSD. In spite of finding four abnormal IIH-associated radiographic variables in certain patients, only 40% of these demonstrated papilloedema.

Our study suggests that the relationship between radiological variables and intracranial pressure (ICP) is complex. To better understand this relationship, we used multivariate modelling methods, which allowed us to make a more precise characterisation. By using continuous values of ICP, we eliminated any binary assumptions about what defines raised ICP. We found that three markers (Yuh sella grade, VT, ODP) remained positively associated with ICP even after adjusting for other clinico-radiological variables, and our model was able to predict median ICP readings with low error (±2.83mmHg). This allowed us to quantify the influence of individual biomarkers on median 24-hour ICP readings.

Whereas many are moving towards building flexible, high-capacity models which can learn complex interactions between multiple imaging predictors, this approach is not easily applied in this specialised domain -owing to the low numbers of patients with complex neurosurgical diagnoses. Given their resistance to overfitting, Bayesian methods are highly appropriate, and demonstrate an improved model fit when compared to classical techniques. We derive a compact, robust regression model, that is transparent and straightforward to interpret by non-expert users, with a clinically useful cross-validated error rate (± 2.71mmHg) using routinely acquired neuroimaging biomarkers that can be incorporated into practice outside of specialised centres. We believe this supports the generalisability of our findings to other similar patient cohorts with diverse CSF dynamic disorders.

Our study also found that the pituitary gland is specifically responsive to attempted ICP normalization following permanent CSF diversion. Although large prospective studies investigating radiographic reversibility following neurosurgical intervention are limited, our results are consistent with previous studies following lumbar puncture [21] or medical treatment [20]. It is possible that markers such as ONSD may have also shown significant reversal if adequately powered or with greater interval time. The clinical utility of this reversibility is as yet unclear, although it could potentially signify successful treatment in patients alongside direct ICP measurement. However, the inclusion of only 36 patients in this specific subgroup analysis reduces the certainty of our findings.

### Limitations and strengths

We note several other limitations in our work. Firstly, this was a retrospective study design that included mostly elective admissions for investigating sub-acute CSF dynamic disorders, making our findings less generalizable to patients who present emergently or have other conditions. Secondly, there was a latency period between MRI scan and ICP monitoring, in which there could be radiographic changes and reduce the reliability of our principal findings. However, this variable did not seem to affect the model fit or the significance of the radiological predictors when incorporated into the multivariate model (Supplementary Results). Thirdly, some radiological variables reported in the literature, such as venous sinus stenosis [21] and other sequences [10,26], were not recorded due to our institutional protocol. Moreover, other outcome variables, such as intracranial pressure amplitude, could have been of interest, but outside the scope of this work. In spite of these limitations, we highlight certain aspects of our study design, namely the use of both frequentist and Bayesian methods, the size of the patient cohort and the diagnostic diversity within it as key strengths which facilitate robust, generalisable results to be obtained, that rely on only simple clinico-radiological parameters and conventionally acquired MRI data.

## Conclusion

In conclusion, our study found significant and independent positive associations between three conventionally acquired MRI biomarkers: Yuh sella grading, vertical tortuosity and optic disc protrusion, and 24-hour median intracranial pressure (ICP), following multivariate adjustment. These results were obtained using both frequentist and Bayesian approaches and were robustly demonstrated by a compact Bayesian multiple linear regression model. Future studies with a prospective design would be strengthened by incorporating a standardised imaging protocol optimised for the assessment of ophthalmic and sella structures. Overall, our study offers a practical tool for screening, triaging, and referring patients with abnormal ICP with greater precision, thereby facilitating clinical decision-making.

## Supporting information

TRIPOD checklist

## Data Availability

All data produced in the present study are available upon reasonable request to the authors, providing permissions have been granted by the ethics committee

## Contributions

A.S.P. and M.C. designed and conceptualised the study

A.S.P, M.C, R.J, A.H.B.J, S.B.J, C.S.T, Z.A., Y.A., M.A., P.D.W.B., M.K.S., E.B.C.T, F.C.Y and H.A. collected and curated the data

A.S.P., M.C., Z.A., Y.A., M.A. and E.B.C.T. performed exploratory data analysis

A.S.P performed statistical modelling

A.S.P., A.K.T. and J.K.R performed validation of imaging biomarkers

A.S.P. and M.C. wrote the first draft of the manuscript All authors edited and revised the manuscript

P.N. and A.K.T. supervised the study

