## Supplementary Material for "The utility of MRI radiological biomarkers in determining intracranial pressure"

### Supplementary Methods

#### Neuroradiology: biomarker assessment

First, an initial subsample of patients with exemplar images were verified and graded. These were reported by a board-certified neuroradiologist and their grading confirmed by an adult hydrocephalus neurosurgical consultant (A.K.T). Second, these images were used as an educational training set for a cohort of medical and clinical neuroscience trainees, who were initially tested to ensure quality assurance and then permitted to grade the remainder of the sample using a crowd-based annotation approach [[1]](https://app.readcube.com/library/829d0e09-047f-41e4-a790-84f30ccd2829/all?uuid=28644486081783527&item_ids=829d0e09-047f-41e4-a790-84f30ccd2829:cb6a4118-d7ac-4157-a502-4a5d4ddd8311). Each patient’s scans were independently scored by two reviewers, masked to the results of their ICP monitoring results and clinical diagnosis. Any discrepancies between reviewers’ scores were resolved with mediation by a senior neurosurgical resident (A.S.P) with an extensive background in neuroimaging. Finally, gradings were independently assessed by a board-certified neuroradiologist (J.R.) using a randomised 1 in 4 subsample. For this data, inter-rater reliability for continuous data (Yuh) was assessed using the intra-class coefficient with a result of 0.85, indicating ‘excellent’ inter-rater agreement [[2]](https://app.readcube.com/library/829d0e09-047f-41e4-a790-84f30ccd2829/all?uuid=8914756620518782&item_ids=829d0e09-047f-41e4-a790-84f30ccd2829:742b65bb-1335-48a3-9e10-135a9ddb1d12). Weighted Cohen’s Kappa [[3]](https://app.readcube.com/library/829d0e09-047f-41e4-a790-84f30ccd2829/all?uuid=9937852605483994&item_ids=829d0e09-047f-41e4-a790-84f30ccd2829:2b44cdee-5ee2-4001-91bf-3d37cc0e5165) was used to assess ordinal data, with ‘substantial’ inter-rater agreement found for globe flattening (0.75) and optic disc protrusion (0.69) and ‘moderate’ agreement for vertical tortuosity (0.46) and optic nerve sheath distension (0.41).

#### Data analysis

Here, weakly informative priors were automatically generated for each explanatory variable to help provide slight regularisation and stabilise computation [[4]](https://app.readcube.com/library/829d0e09-047f-41e4-a790-84f30ccd2829/all?uuid=22585010648219161&item_ids=829d0e09-047f-41e4-a790-84f30ccd2829:111b02c2-18de-4dab-87e3-9263b5be42d8), and an adaptive Hamiltonian Monte Carlo algorithm [[5]](https://app.readcube.com/library/829d0e09-047f-41e4-a790-84f30ccd2829/all?uuid=02585788562366742&item_ids=829d0e09-047f-41e4-a790-84f30ccd2829:0a465ab4-c991-4d59-b906-fece021a4327) is used to sample from the joint posterior distribution of the parameters (with 10000 draws per chain and 5000 tuning burn-in). Model convergence and diagnostics of both priors and posterior distributions were visually inspected. Both quadratic terms and scaling independent variables were assessed and models compared using the Watanabe-Akaike Information Criterion.

Patients with missing data were removed from the model, and no imputation was used. Study size was determined pragmatically using all the available data. However, post-hoc analysis using the weakest model (OLS radiological, multivariate) with an adjusted R2 of 0.26 would yield a Cohen f^2^ effect size of 0.35. With an alpha error of 0.05, and given the number of subjects in the model, this would yield a power of 1 (GPower, v=3.1).

### Supplementary Results

#### Reversibility

**Supplementary Table 1.** Assessment of biomarker reversibility following CSF diversion. (ONSD = optic nerve sheath distension; ODP = optic disc protrusion; PGF = posterior globe flattening; VT = vertical tortuosity; Yuh = pituitary sella grade)

| **Radiological biomarker** | **Pre-intervention:**  **median (IQR)** | **Post-intervention:**  **median (IQR)** | **p**  **(unadjusted)** |
| --- | --- | --- | --- |
| Yuh | 2 (1-3) | 1 (1-2) | 0.0001 |
| VT | 0 (0-1) | 0 (0-1) | 0.23 |
| ONSD | 0 (0-0) | 0 (0-0) | 0.10 |
| PGF | 0 (0-0) | 0 (0-0) | 0.18 |
| ODP | 0 (0-0) | 0 (0-0) | 0.18 |

#### Multivariate prediction of ICP using radiological markers

**Supplementary Figure 1**. A) Boxplot demonstrating ranges of ICP in patients who had abnormal radiological biomarkers. B) Cramer’s V heatmap demonstrating the degree of co-linearity across radiological variables

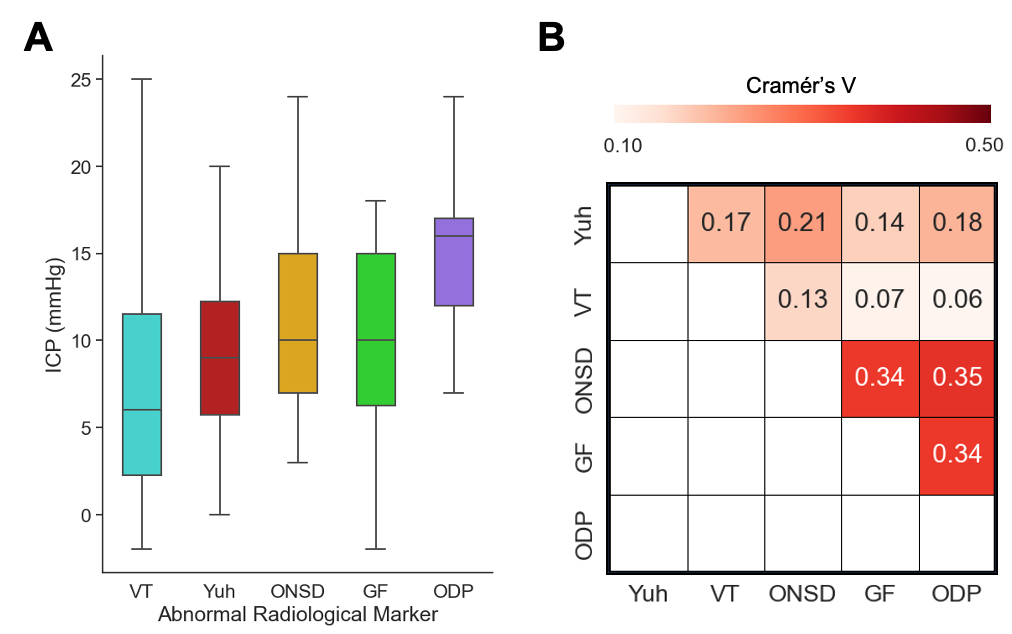

**Supplementary Table 2.** Multivariate ordinary least square linear regression model using clinico-radiological parameters to predict ICP. IIH = idiopathic intracranial hypertension; NPH = normal pressure hydrocephalus; LOVA = long standing overt ventriculomegaly in adults; ODP = optic disc protrusion, PGF = posterior globe flattening; Yuh = pituitary sella grade; VT = vertical tortuosity

| **Variable type** | **Variable** | **Coefficient** | **t** | **p** |
| --- | --- | --- | --- | --- |
| Intercept | | 3.5089 | 1.911 | 0.057 |
| **Clinical** | ICPAge | -0.0700 | -3.030 | 0.003 |
|  | Aqueductal stenosis | Reference |  |  |
|  | Congenital | -1.2101 | -0.578 | 0.564 |
|  | Chiari-syringomyelia | -1.0103 | -0.589 | 0.556 |
|  | IIH | 2.4758 | 1.488 | 0.138 |
|  | CSF Leak | -2.9163 | -1.161 | 0.247 |
|  | LOVA | 1.8015 | 0.744 | 0.457 |
|  | NPH | -0.4743 | -0.216 | 0.829 |
|  | Secondary following neurovascular injury | 5.2441 | 1.451 | 0.148 |
|  | Other | -1.0654 | -0.618 | 0.537 |
|  | TBI | -4.2658 | -1.187 | 0.236 |
|  | Secondary following tumour | 0.1697 | 0.092 | 0.927 |
| **Radiological** | Yuh | 1.7566 | 6.094 | 0.000 |
|  | ODP | 3.6221 | 2.967 | 0.003 |
|  | VT | 1.9561 | 3.990 | 0.000 |
|  | GF | 0.6917 | 0.830 | 0.407 |

With the addition of the time interval between MRI and ICP monitoring in the multivariate OLS model, adjusted R^2^ marginally reduced to 0.32, and the three radiographic variables remained significant (Yuh sella grade, coef = 1.76, p = <0.0001; ODP, coef = 3.70, p = 0.003; VT = 1.89, p = < 0.0001).

**Supplementary Table 3.** Bayesian linear regression model using clinico-radiological parameters to predict ICP with model parameters. (IIH = idiopathic intracranial hypertension; NPH = normal pressure hydrocephalus; LOVA = long standing overt ventriculomegaly in adults; ODP = optic disc protrusion, PGF = posterior globe flattening; Yuh = pituitary sella grade; VT = vertical tortuosity)

| **Variable type** | **Variable** | **Mean**  **Coefficient** | **SD** | **3% HDI** | **97% HDI** |
| --- | --- | --- | --- | --- | --- |
| Intercept | | 3.491 | 1.855 | -0.030 | 6.949 |
| **Clinical** | ICPAge | -0.070 | 0.023 | -0.114 | -0.026 |
|  | Aqueductal stenosis | Reference |  |  |  |
|  | Congenital | -1.204 | 2.116 | -5.170 | 2.792 |
|  | Chiari-syringomyelia | -1.001 | 1.729 | -4.277 | 2.224 |
|  | IIH | 2.486 | 1.681 | -0.750 | 5.557 |
|  | CSF Leak | -2.905 | 2.524 | -7.647 | 1.855 |
|  | LOVA | 1.821 | 2.451 | -2.647 | 6.518 |
|  | NPH | -0.469 | 2.204 | -4.800 | 3.531 |
|  | Secondary following neurovascular injury | 5.255 | 3.684 | -1.730 | 12.108 |
|  | Other | -1.064 | 1.741 | -4.353 | 2.195 |
|  | TBI | -4.265 | 3.637 | -11.062 | 2.451 |
|  | Secondary following tumour | 0.182 | 1.866 | -3.220 | 3.778 |
| **Radiological** | Yuh | 1.756 | 0.289 | 1.191 | 2.283 |
|  | ODP | 3.612 | 1.220 | 1.328 | 5.891 |
|  | VT | 1.954 | 0.490 | 1.051 | 2.879 |
|  | GF | 0.699 | 0.836 | -0.832 | 2.320 |
| Regression parameters:  Median ICP sigma (SD) = 5.58 mmHg (0.23)  Family = Gaussian  Weakly informative priors scaled and normally distributed with mu = 0, and scaled standard deviations based on patient sample | | | | | |
